# Assessment of Knowledge on Human Mpox Virus among General Population in Bangladesh

**DOI:** 10.1101/2022.08.31.22279445

**Authors:** Sudipta Deb Nath, A.M. Khairul Islam, Koushik Majumder, Fahmida Hoque Rimti, Jyoti Das, Mustari Nailah Tabassum, Arefin Naher Oishee, Tarannum Mahmood, Monisha Paul, Muntasrina Akhter, Alok Bijoy Bhadra, Fariha Hoque Rimu, Snahasish Chakraborty, Preetom Shom, Sirajum Monira Nosaibah, Md Ashikur Rahman, Ahmed Safwan Khan, Anika Anjum, Sushmita Khan, M. Mahbub Hossain, Mohammad Delwer Hossain Hawlader

## Abstract

**Introduction:** Mpox (Monkeypox) is a zoonotic Orthopox virus of the Poxviridae family. The general public in Bangladesh should be informed about Mpox to reduce the burden of a possible epidemic in the community.

**Objective:** The study aimed to determine the level of public awareness and knowledge about Mpox among the general community to provide information regarding future preventive measures.

**Materials and Method:** From May to June 2022, this nationwide cross-sectional study was conducted in eight administrative divisions in Bangladesh. We determined the sample size using Cochran’s formula. Through a semi-structured questionnaire, data regarding sociodemographic characteristics and knowledge about Mpox were collected via face-to-face interviews. IBM SPSS v.25 was used to analyze the data.

**Result:** Among the total of 1,711 respondents to the questionnaire, almost two-thirds (N=1139) of the respondents had heard about Mpox prior to the study. We observed poor knowledge scores (mean ± sd) about the transmission pathways (0.71 ± 0.73), vaccination (0.09 ± 0.27), and the signs and symptoms of Mpox (1.91 ± 1.50). Most participants were also unaware of the treatment options of Mpox (0.22 ± 0.59). Educational status and occupation were found to affect the knowledge significantly (p value<0.001). This study showed that the higher the education level, the higher the knowledge level.

**Conclusion:** The general community has a minimal understanding of the spread of Mpox and its prevention. This virus requires additional research on its epidemiology, ecology, and biology in endemic regions to be comprehended and prevented.

## 1. Introduction

Mpox (Monkeypox) is a zoonotic disease caused by infection with the Mpox virus, a member of the Orthopox virus genus (1,2). The virus was first detected in 1958 in the Cynomolgus monkey colony (1). Afterward, in 1971, the first Mpox case was identified in the Democratic Republic of Congo [DRC], and following this, the disease became endemic in central and West Africa (2–4). However, the first incidence outside Africa occurred in the United States in 2003, caused by a rodent imported from Ghana (1,2,4). The detection of cases in the UK and Israel in 2018 and in Singapore in 2019 was documented accordingly (5,6). More than 86,000 cases have been reported in 113 nations, prompting the World Health Organization (WHO) to declare this a public health emergency of international significance (7). This new emergence has more variations compared to its previous presentations such as younger patients, higher prevalence in men, higher rates of sexual transmission, anogenital lesions, an incubation period of up to 21 days, and infection without traveling to an endemic area (8–10). Moreover, this disease has similar presentations to smallpox including fever accompanied by headache, rash, back pain, malaise, and fatigue, except for the specific symptom of lymphadenopathy (1,2).

The virus is transmitted through bushmeat handling, animal hosts, such as squirrels, rodents, and prairie dogs, infected oropharyngeal secretion, and direct contact (1,2,11). Several risk factors, such as the interaction between humans and infected animals, termination of smallpox vaccination, and increased globalization, make the disease a future global public health concern (1,12). Also, the rising occurrence among the younger unprotected population raises concerns about the imminent spread of the human Mpox virus (13).

Although the reason for re-emergence is still unknown, its broad range of animal hosts with high adaptation power and ability to circulate among humans is already making the virus alarming for the future (14). A report by WHO revealed that a lack of information about the Mpox was one of the obstacles in preventing its re-emergence (15). Studies done on general practitioners revealed that only 10% (Indonesia), 18.6% (Saudi Arabia), and 27% (Italy) of the participants had sufficient knowledge about Mpox (16–18). Another study in Saudi Arabia showed that 48% of the general population had sufficient knowledge about Mpox (19). To limit the potential for the Mpox virus spreading across the world, the general public must be educated about the disease and know how to protect themselves. Additionally, due to the dense population, disease transmission is facilitated in Bangladesh (20). Thus, this study aims to measure the level of awareness and knowledge about Mpox among the general population of Bangladesh, in order to guide future prevention and treatment planning.

## 2. Methodology

### 2.1. Study design, settings, and ethics

This was an observational type of cross-sectional study and comprised participants from all the eight divisions (Barishal, Chattogram, Dhaka, Khulna, Mymensingh, Rajshahi, Rangpur, and Sylhet) of Bangladesh to provide a nationwide representation. The survey was conducted from May to August 2022, immediately after the confirmation of the ongoing global outbreak of the human Mpox virus in 2022. We followed the simple random sampling method to collect our data. Informed consent was collected from every participant. The anonymity and confidentiality were ensured by excluding the name of the participants. Ethical clearance for the study was taken from the Institutional Review Board (IRB) of North South University.

### 2.2. Criteria of inclusion and exclusion

The criteria of inclusion for participants were 1) being a Bangladeshi resident, 2) being an adult (≥ 18=years old), and 3) providing consent. The exclusion criteria included-1) having any psychiatric illnesses, and 2) not providing consent.

### 2.3. Sample size

Considering an unknown prevalence (50%), 5% margin of error, 95% confidence interval, and 165,158,616 population size a minimum sample size was calculated of 383 (21). However, to increase the diversity and representativeness of the study sample, efforts were made to enrol the maximum number of participants, therefore a final sample size of 1743 was achieved.

### 2.4. Development and validation of the survey questionnaire

A 20-item semi-structured questionnaire was first developed following Center for Disease Control and Prevention (CDC) guidelines and a study by Harapan H. *et al.* (15,16,22). Then, a pilot study was undertaken on 85 participants to validate the questionnaire. The validation method of the questionnaire included conducting two interviews with the same 85 participants. The second interview was conducted one week after the first one to assess whether the participants were able to comprehend the questions consistently across both sessions. Finally, the questionnaire was revised, upgraded, and divided into two sections to collect data (Supplementary File 1). The first section consisted of sociodemographic variables, such as gender, age, education, and profession. The second section consisted of 12 questions regarding knowledge of the Mpox outbreak, its signs, symptoms, prevention, and treatment. Moreover, we translated the questionnaire into Bengali for easy understanding.

### 2.5. Data Collection

All the data was collected via face-to-face interviews. Ten data collectors collected data after one week of training regarding the data collection tools and measures for ensuring optimal participation. It took approximately 10 and 12 minutes to complete the whole questionnaire. The objective of this study was described to all the participants in their native language (Bengali). In addition, including the pilot-study participants, we collected 101 data from Barishal, 309 from Chattogram, 286 from Dhaka, 279 from Khulna, 213 from Mymensingh, 176 from Rajshahi, 219 from Rangpur, and 213 from Sylhet.

### 2.6. Study variables

In our study, we observed the percentage of persons knowing/hearing the name of Mpox, which is termed as ‘awareness’. The known details about the disease is termed as ‘knowledge’ (23). Knowledge-based questions were for those who at least knew/heard of the name ‘Mpox’. For the questions regarding knowledge of Mpox, participants were given 1 for each correct answer and 0 for each incorrect/ do not know ones. For the questions with more than one correct answer, each correct option was given 1 point. However, only for question 12 in Table 3, the scoring was −2 to +2. The highest knowledge score was 26. The mean score per question is demonstrated in Table 3, and the mean percent knowledge score (total knowledge score * 100 / 26) difference concerning sociodemographic variable are in Table 4. We also collected the age (18-35 years/ 36-50 years/ above 50 years), gender (male/ female), education (no formal education/ primary/ secondary/ higher secondary/ graduate/ postgraduate), occupation (unemployed or retired/ student/ homemaker/ daily worker/ service holder/ business/ health professionals), and residence (rural/ urban) as sociodemographic variables.

### 2.7. Statistical analysis

All the data analysis was done using IBM SPSS v.25. The illustration was prepared by GraphPad Prism (version 8.0). Descriptive analysis of the sociodemographic variables is shown in Table 1. Simple and multiple binary logistic regression was used to examine the odds of demographic variables on being aware of Mpox. Omnibus Tests of Model Coefficients and Hosmer and Lemeshow Goodness of Fit Test were performed to see the validity of the regression model. Results were presented as crude and adjusted odds ratios. Independent-Samples T-test and One-way ANOVA test were conducted to observe the mean knowledge-score difference among the categories of the sociodemographic information. P values less than 0.05 were considered significant.

**Table 1:** Sociodemographic information of the participants.

| Sociodemographic information | Category | Number (%) |
| --- | --- | --- |
| Age (in years) (N = 1711) | 18-35 | 1102 (64.4%) |
|  | 36-50 | 349 (20.4%) |
|  | Above 50 | 260 (15.2%) |
| Gender (N = 1711) | Male | 979 (57.2%) |
|  | Female | 732 (42.8%) |
| Educational Status (N = 1711) | No formal education | 142 (8.3%) |
|  | Primary | 107 (6.3%) |
|  | Secondary | 199 (11.6%) |
|  | Higher secondary | 346 (20.2%) |
|  | Graduate | 664 (38.8%) |
|  | Postgraduate | 253 (14.8%) |
| Occupation (N = 1711) | Unemployed/retired | 97 (5.7%) |
|  | Student | 593 (34.7%) |
|  | Homemaker | 197 (11.5%) |
|  | Daily worker | 187 (10.9%) |
|  | Service holder | 307 (17.9%) |
|  | Business | 212 (12.4%) |
|  | Health professionals | 118 (6.9%) |
| Residence (N = 1711) | Rural | 427 (25.0%) |
|  | Urban | 1284 (75.0%) |

## 3. Results

### 3.1. Sociodemographic Information

In total, 1711 respondents completed the questionnaire. Male (N=979/1711; 57.2%) made up most of the cohort. More than one-third of the respondents (N=1102/1711; 64.4 %) were between 18 and 35, and more than one-third indicated a graduate degree as their highest educational attainment (N=664/1711; 38.8 %). Three-fourths of the participants reported living in an urban area (N=1284/1711; 75.0%), as shown in Table 1.

### 3.2. Sources of knowledge about Mpox

About 61.7% of the respondents (N=1055/1711) were aware of Mpox, while 38.3% (N=656/1711) were unaware of such virus (Figure 1A). The respondents who were aware of Mpox also reported the source from which they obtained Human Mpox virus-related information. Most of the participants acquired their information from social media (N = 508/1055; 48.2%) and the internet (N= 355; 33.6%) followed by Television (340/1055; 32.2%), Newspaper (179/1055; 17.0%), friends/neighbours (148/1055; 14.0%), and family (60/1055; 5.7%) (Figure 1B). On the contrary, healthcare personnel (N=16/1055; 1.5%) were the least influential source of information for the participants (Figure 1B).

**Figure 1:**
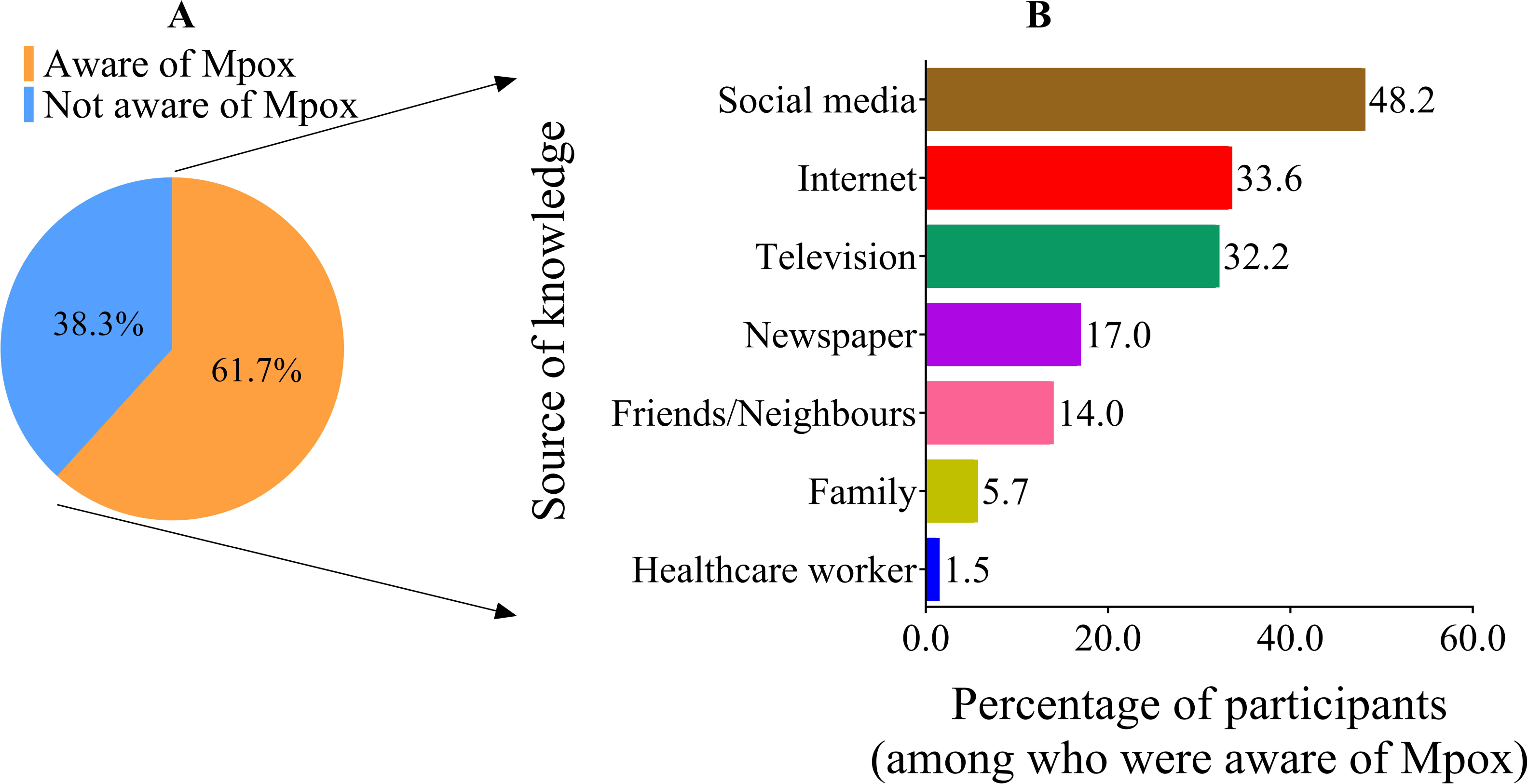
Awareness of the respondents about Mpox and their source of knowledge. In Figure 1A, the pie chart shows if respondents were previously aware of Mpox or not. In Figure 1B, the bar chart shows the sources of knowledge about Mpox, where the X-axis denotes the percentage of participants (of those with awareness about Mpox) and the Y-axis shows the sources.

### 3.3. Awareness about Mpox

Gender, occupation, and place of residence had statistically significant effects on respondents’ awareness of Mpox (Table 2). Gender of the participants was associated with their awareness of monkeypox, where males were more aware than females (AOR: 1.4, 95% CI [1-1.8], p<0.001). Respondents having secondary level (AOR: 4, 95% CI [2.1-7.4], p<0.001), higher secondary level (AOR: 8.1, 95% CI [4.2-15.4], p<0.001), graduate level (AOR: 11.2, 95% CI [5.9-21.4], p<0.001) and post-graduate level (AOR: 9.8, 95% CI [5.0-19.2], p<0.001) education was significantly associated with awareness and the odds of respondents being aware of this virus increased with their higher education level. Respondents from urban areas were more likely to be aware of Mpox (AOR: 1.4, 95% CI [1.7-2.3], p<0.001). (Table 2)

**Table 2:** Factors affecting the awareness of the respondents about Mpox.

| Variables | Crude odds ratio (COR) | Range of COR (95% CI) | P-value | Adjusted odds ratio (AOR) | Range of AOR (95% CI) | P-value |
| --- | --- | --- | --- | --- | --- | --- |
| Age (in years) |  |  |  |  |  |  |
| 18-35 | 2.7 | 2.0-3.5 | <0.001* | 1.3 | 0.9-1.9 | 0.118 |
| 36-50 | 1.2 | 0.9-1.7 | 0.193 | 1.3 | 0.9-1.9 | 0.207 |
| Above 50 (Ref) | 1.0 | - | - | 1.0 | - | - |
| Gender |  |  |  |  |  |  |
| Male | 1.0 | 0.8-1.2 | 0.972 | 1.4 | 1-1.8 | 0.021* |
| Female (Ref) | 1.0 | - | - | 1.0 | - | - |
| Educational Status |  |  |  |  |  |  |
| No formal education (Ref) | 1.0 | - | - | 1.0 | - | - |
| Primary | 3.0 | 1.6-5.8 | 0.001* | 2.6 | 1.3-5.1 | 0.005* |
| Secondary | 5.7 | 3.2-10.1 | <0.001* | 4.0 | 2.1-7.4 | <0.001* |
| Higher secondary | 15.4 | 8.8-26.7 | <0.001* | 8.1 | 4.2-15.4 | <0.001* |
| Graduate | 23.4 | 13.7-40.0 | <0.001* | 11.2 | 5.9-21.4 | <0.001* |
| Post-graduate | 18.5 | 10.4-32.9 | <0.001* | 9.8 | 5.0-19.2 | <0.001* |
| Occupation |  |  |  |  |  |  |
| Unemployed/retired (Ref) | 1.0 | - | - | 1.0 | - | - |
| Student | 2.1 | 1.3-3.2 | 0.002* | 1.2 | 0.7-2 | 0.533 |
| Homemaker | 0.7 | 0.4-1.1 | 0.090 | 0.9 | 0.5-1.7 | 0.772 |
| Daily worker | 0.2 | 0.1-0.3 | <0.001* | 0.6 | 0.3-1.1 | 0.094 |
| Service holder | 1.6 | 1.0-2.6 | 0.040* | 1.1 | 0.6-1.8 | 0.829 |
| Business | 0.8 | 0.5-1.3 | 0.371 | 0.9 | 0.5-1.6 | 0.852 |
| Health professionals | 2.5 | 1.3-4.5 | 0.003* | 1.4 | 0.7-2.6 | 0.337 |
| Residence |  |  |  |  |  |  |
| Rural (Ref) | 1.0 | - | - | 1.0 | - | - |
| Urban | 2.8 | 2.2-3.5 | <0.001* | 1.7 | 1.4-2.3 | <0.001* |

\*P-value significant; CI: Confidence Interval.

### 3.4 Knowledge about Mpox

A 12-item questionnaire was used to assess knowledge about Mpox among those who were aware of the virus. The distribution of each knowledge item about Mpox and the mean scores against each question are presented in Table 3

**Table 3:** Knowledge score assessment of Mpox by individual questions.

| Knowledge assessment | Scoring (mean ± sd) |
| --- | --- |
| 1. Knowledge of Mpox prevalence in Africa. (0-1) | 0.66 ± 0.47 |
| 2. Knowledge regarding the outbreak of human Mpox in Europe, the USA, UK. (0-1) | 0.61 ± 0.49 |
| 3. Knowledge of Human Mpox cases in Bangladesh. (0-1) | 0.57 ± 0.50 |
| 4. Knowledge of Mpox as a viral disease. (0-1) | 0.68 ± 0.47 |
| 5. Knowledge regarding carrier of Mpox virus (squirrels, rat/mice, monkey). (0-3) | 0.71 ± 0.73 |
| 6. Knowledge of signs and symptoms of Mpox. (0-5) | 1.91 ± 1.50 |
| 7. Knowledge of similar signs and symptoms of Mpox and smallpox. (0-1) | 0.48 ± 0.50 |
| 8. Knowledge on the transmission of Mpox. (0-5) | 0.86 ± 1.12 |
| 9. Knowledge on international travellers as a source of imported cases of Mpox. (0-1) | 0.81 ± 0.39 |
| 10. Knowledge of prevention of Mpox virus infection. (0-4) | $2.00 \pm 1.53$ |
| 11. Knowledge on vaccination against the Mpox virus. (0-1) | $0.09 \pm 0.27$ |
| 12. Knowledge regarding the treatment of Mpox virus infection. (-2 - +2) | $0.22 \pm 0.59$ |

Most of the respondents answered correctly (0.66 ± 0.47) identifying Africa as the most prevalent region for Mpox. Similarly, the mean score was also 0.61 ± 0.49 about the recent outbreaks of Mpox in Europe, the USA, and the UK. In regard to any cases of Mpox in Bangladesh, knowledge of the respondents about this item was 0.57 ± 0.50. The majority of the participants could identify Mpox as a viral disease, thus the scoring was 0.68 ± 0.47. In response to the question about the source of Mpox, participants were able to choose one or more options from squirrels, rat/mice, and monkeys. As all these animals may be responsible for spreading Mpox, the mean scoring of this item (0.71 ± 0.73) indicates that respondents are largely unaware of the sources of Mpox (Table 3).

About signs and symptoms of Mpox, participants could select one or more of the following: fever and headache, rash, muscle discomfort, swollen lymph node, and skin lesion. The mean score (1.91 ± 1.50) was lower in this issue (Table 3). Most of the participants were unable to correctly identify that Mpox and smallpox do indeed have similar symptoms (0.48 ± 0.50). About the mode of transmission, participants could select one or more alternatives from an animal bite or scratch, wild animal meat preparation/processing, significant respiratory, indirect contact with lesion material, and mother to fetus. We found that the knowledge level on this matter was very little (0.86 ± 1.12). The scoring about international travellers as a source of imported cases of Mpox was 0.81 (± 0.39), indicating that most of the respondents were aware on this regard (Table 3).

Regarding how they would prevent Mpox, participants were given the option of avoiding contact with sick animals that could harbor the virus, isolating infected people from others, avoiding direct contact with infected materials, or practicing good hand hygiene after contact with infected animals or humans. As all of these are prevention methods of Mpox, the mean scoring of this item (2.00 ± 1.53) indicates that knowledge level about the transmission pathways was lower. Almost all respondents were unaware of a vaccine (0.09 ± 0.27). Additionally, most of the participants were unable to correctly identify the treatments for Mpox (0.22 ± 0.59) (Table 3).

### 3.5. Factors that influence knowledge scores

In Table 4, the factors affecting the knowledge score were demonstrated. According to the findings, individuals with a higher degree of education have considerably higher mean knowledge scores about Mpox, and the higher the education level, the higher the knowledge level was.

**Table 4:** Factors affecting knowledge score regarding Mpox.

| Variables | Knowledge percent score | P-value |
| --- | --- | --- |
| Age (in years) |  |  |
| 18-35 | 37.22 ± 19.69 | 0.676 |
| 36-50 | 36.66 ± 20.45 |  |
| Above 50 | 35.53 ± 20.13 |  |
| Gender |  |  |
| Male | 36.57 ± 20.14 | 0.485 |
| Female | 37.43 ± 19.49 |  |
| Educational Status |  |  |
| No formal education | 16.74 ± 18.14 | <0.001 |
| Primary | 25.56 ± 17.44 |  |
| Secondary | 28.91 ± 19.78 |  |
| Higher secondary | 35.72 ± 19.96 |  |
| Graduate | 38.79 ± 19.39 |  |
| Post-graduate | 41.05 ± 18.80 |  |
| Occupation |  |  |
| Unemployed/retired | 36.98 ± 19.43 | <0.001 |
| Student | 35.55 ± 19.63 |  |
| Homemaker | 29.23 ± 17.40 |  |
| Daily worker | 24.86 ± 19.04 |  |
| Service holder | 41.52 ± 18.40 |  |
| Business | 32.68 ± 19.88 |  |
| Health professionals | 51.46 ± 17.05 |  |
| Residence |  |  |
| Rural | 36.37 ± 20.85 | 0.683 |
| Urban | 37.06 ± 19.65 |  |

*\*P-value significant*

Respondents with a primary level of education had a higher mean percent knowledge score (25.56 ± 17.44) than those without formal education (16.74 ± 18.14). The mean knowledge score of respondents with a higher secondary degree (35.72 ± 19.96) was greater than that of respondents with a secondary degree (28.91 ± 19.78) (Table 4). A higher mean knowledge score (41.05 ± 18.80) was also obtained by respondents with post-graduate degrees compared to those with graduate degrees (38.79 ± 19.39). Education and occupational factors significantly influenced the mean knowledge score (p<0.001). Among all the occupations, healthcare professionals had the highest mean knowledge score (51.46 ± 17.05).

## 4. Discussion

The results of the study indicated that the general people lacked essential awareness and understanding about Mpox. About two-thirds of the respondents heard about Mpox, while 38.3% had never heard of the name-‘Mpox’ before completing the survey. In a Malaysian study, researchers indicated that almost 95% of respondents were aware of dengue fever, another viral illness (which is also more common), during its epidemic, despite their lack of understanding of transmission and management (24). A pilot study in Karachi, Pakistan, was conducted among people that visited hospitals. The findings indicated that 90% of the participants were aware of the viral disease, among whom only 38.5% possessed adequate knowledge (25). Similarly, another survey from Pakistan indicated that only 35% of the adults from low and high socioeconomics groups possessed sufficient knowledge of dengue during the outbreak (26). Given that dengue is more prevalent than monkeypox, it is not surprising for our participants to be unfamiliar with the name Mpox. Further investigation is needed in order to comprehend how to better inform the general population about a newly discovered, potentially developing virus (27,28).

The news of potential Human Mpox virus cases in Bangladesh spread like wildfire on multiple social media channels. On the other hand, the Bangabandhu Sheikh Mujib Medical University (BSMMU) refuted these assertions, stating that no case of Mpox had yet been identified (29). Given the global discovery of the virus, there is an urgent need to understand the source better, the dynamics of transmission, and to give people the knowledge and assistance they require to safeguard themselves and others in various situations (30). To the best of our knowledge, this is the first nationwide survey in Bangladesh to look into the awareness of general public and understanding of the fundamentals, spread, transmission, symptoms, prevention, and treatment of Human Mpox virus infection.

Most research participants were generally ignorant of the origins of the virus. Moreover, the degree of understanding regarding the transmission channels is also insufficient with only 0.86 mean score according to our findings, while the maximum score for the question was five. A WHO study indicated that one of the difficulties in avoiding the re-emergence of Mpox was a lack of information about Mpox, which is supported by our findings (30). Although the majority of participants in the survey could not identify that Mpox and smallpox have comparable symptoms, almost all of the individuals identified that Mpox is a viral illness that affects them. Vaccination is one of the best ways to prevent infectious illnesses. As there are a few effective antiviral medications for Mpox, in some high-risk circumstances, vaccines to prevent the human smallpox infection (offering 95% Mpox protection), might be utilized (31,32). As per our findings, most of the respondents were unaware about the vaccine and unable to correctly name the treatments as well. Nevertheless, owing to a lack of information, the general population of Bangladesh is mostly in the dark about the identity and diversity of viruses prevalent worldwide.

We also demonstrated that those who use the internet or social media as a source of information are more knowledgeable than their peers about Mpox. This might be because social media and the internet make it simple for most individuals to access information that has been updated. This emphasizes the usefulness of the internet for health promotion, especially in the case of pandemics (33). Compared to other resources, online media have emerged as one of the primary and expedient sources for information access (34). These findings are similar to other studies performed in Egypt and Ethiopia that mentioned the internet and social media as the main information sources (35,36). However, the advent of social media has also accelerated the widespread dissemination of inaccurate data which can be challenging to detect. On top of that its ability to impact the cognition and convictions of individuals is evident (37). Newspapers, local leaders, and healthcare professionals appear to be a less common source of knowledge regarding the ailment when compared to other media. As a result, the Bangladeshi health system may increase the involvement of community leaders and health extension workers to disseminate information to the populace efficiently.

In line with the majority of previous research done in developing countries (38), we observed that men had a higher awareness of this viral illness than females in our study. In developing countries like Bangladesh, rural males usually have more outside interactions than females. This might have affected the overall gender-based knowledge differences in this study. Consistent with previous research (39,40), we found that as the education level of the participants grew, so did their awareness about the spread of this virus. Furthermore, the knowledge of people from the urban area was 1.4% higher than that of their rural counterparts. The outcome of a Chinese study (41) is consistent with this observation, where the knowledge scores for urban and rural residents were 25.58 ± 3.73 and 24.35 ± 3.95, respectively. The greater availability of mass media might explain this in urban areas, such as television, radio, and online social networks (42). While health professionals exhibited the highest knowledge score in this study, there is still room for further improvement through targeted training. In situations of endemics or pandemics, healthcare workers consistently receive guidance to effectively map the outbreak and implement essential interventions aimed at limiting its transmission (43). The knowledge of general public in this field may also be influenced by the availability of suitable facilities to support health educators, including medical personnel and government officials.

The findings of this study may have several implications for public health policymaking and practice. First, the lack of accurate knowledge of Mpox in the Bangladeshi population amidst a global public health crisis requires multipronged interventions that promote education and preventive health behavior. Community-based, social media and mass media interventions have been effective in health promotion among the Bangladeshi population in the past, which informs the potential of these media to promote Human Mpox virus knowledge and prevention (44,45). Second, healthcare providers and social workers need to work with multisectoral organizations to mobilize organizational resources that ensure optimal preparedness for potential Human Mpox virus outbreaks and associated health hazards in Bangladesh. Third, community stakeholders should be engaged in participatory community prevention of Mpox and other infections that may affect individuals and families living in diverse Bangladeshi communities. In this process, exploring and utilizing context-specific knowledge would be critical for successful infection prevention. Fourth, situational analyses at the community, Upazila, district, and divisional level should be conducted in the context of Mpox and how a potential outbreak may impact the current state of healthcare and social services. These insights, alongside other data sources, should be used to create a national-level data dashboard that informs real-time decision-making for the prevention of Human Mpox virus outbreaks and other infection outbreaks. News agencies and other public media can use such a database to generate public knowledge and disseminate the same. Lastly, policy-level efforts should be made proactively to mitigate the potential crises that may arise from an outbreak and lessons learned from COVID-19, and Human Mpox virus outbreaks in similar contexts should be analyzed to inform national and local actions preventing adverse health outcomes in the future.

## Conclusion

This study suggests that the general population of Bangladesh lacks sufficient knowledge about the source of infection and the dynamics of Mpox transmission. Therefore, considering the ongoing virus emergence, it is vital to offer the necessary knowledge and support to help people protect themselves and others. In light of the current global public health crisis, healthcare professionals, public health workers, and healthcare policymakers should collaborate to establish a robust disease surveillance system and implement appropriate policies and interventions that enable effective health education and preventive health behaviors to promote public awareness, increase the precautionary behaviors and mitigate the potential crisis.

## Strengths and Limitations

This is one of the earliest analyses regarding the knowledge of the general people of Bangladesh on Human Mpox virus. As it is a preliminary study and due to limited logistic support, we have been unable to reach out to all the areas in Bangladesh within a short span. However, a larger sample size would provide more extensive and unbiased (narrower confidence intervals with more precision in regression models) insight into the current situation in Bangladesh. Moreover, our data collection plan from each division included considering the corresponding population density of the divisions. However, this aspect could not be pursued due to both time and resource limitations. In future research regarding Mpox, more precise data collection method might be followed.

## Author Contribution

**Sudipta Deb Nath**: Conceptualization, Data analysis, Writing-Original draft preparation, Writing-Reviewing and Editing

**A.M. Khairul Islam**: Data analysis, Writing-Original draft preparation, Writing-Reviewing and Editing

**Koushik Majumder**: Data analysis, Investigation

**Fahmida Hoque Rimti**: Writing-Original draft preparation, Visualization

**Jyoti Das**: Writing-Reviewing and Editing, Data curation

**Mustari Nailah Tabassum**: Writing-Reviewing and Editing

**Arefin Naher Oishee**: Methodology, Data curation

**Tarannum Mahmood**: Data curation, Writing-Original draft preparation

**Monisha Paul**: Data curation, Data analysis

**Muntasrina Akhter**: Methodology, Visualization

**Alok Bijoy Bhadra**: Conceptualization, Validation

**Fariha Hoque Rimu:** Data curation, Writing-Reviewing and Editing

**Snahasish Chakraborty**: Data curation, Investigation

**Preetom Shom**: Data curation, Visualization

**Sirajum Monira Nosaibah**: Methodology, Data curation

**Md Ashikur Rahman**: Data curation, Investigation

**Ahmed Safwan khan**: Conceptualization, Data curation

**Anika Anjum**: Data curation, Visualization

**Sushmita khan**: Data curation, Investigation

**M. Mahbub Hossain**: Methodology, Writing-Reviewing and Editing

**Mohammad Delwer Hossain Hawlader**: Supervision

## Supporting information

Consent form

## Data Availability

URL or Link for data https://osf.io/jkt48/?view_only=5b9c61de4f9148faaa03ce2ec9186cf8&fbclid=IwAR3T5iVBV1cxq8bA82bv4YRuSnwAHvblRyFDFufjJWMh8BcPJe4Mhe43MQ0

---

**Supplementary File 1:** Questionnaire of the study

