## Supplementary material for "Assessment of Knowledge on Human Mpox Virus among General Population in Bangladesh": Consent form

ID/Name: ________________

Dear Participant,

As a part of the research team, I would like to welcome you to our research project on “Assessment of Knowledge on Human Mpox Virus among General Population in Bangladesh.”

This form contains details about this research and will act as a written document for your participation as a research participant. You can consult with us regarding your participation in this research and you can take time to decide.

Mpox is a zoonotic Orthopox virus of the Poxviridae family. The questionnaire for this study is used to determine the public awareness level of Mpox among the general community. We will interview the general population using a semi-structured questionnaire that will take around 10 minutes to complete. Your participation in this research will be completely voluntary and you will not receive any sort of financial or any other benefits. You can withdraw yourself from this research anytime you want.

This research does not include any body fluid or tissue collection, so there is no scope for health hazards. If you are unable to understand any part of the questionnaire you can ask our research volunteer for clarification. All the data collection will be done anonymously, and no information will be published that can identify a participant. The information that we will collect from this research project will be kept confidential and if you wish you can skip any question that seems to be distressful to you. Information that will be collected during the research will be stored in a password-protected electronic format and only the authorized researchers will have access to the data, and it will not be shared with or given to anyone.

If you have any inquiries regarding this questionnaire or the research, you may contact the investigators:

**Dr. A.M.Khairul Islam**

International Centre for Diarrhoeal Disease Research, Bangladesh (ICDDR,b)

**Dr. Sudipta Deb Nath**

Department of Genetic Engineering and Biotechnology, University of Dhaka, Bangladesh

Thank you very much for agreeing to participate in this survey.

**Consent of the participant:**
Name:

Signature: __________________________

**Section-1: Demographic Information**

| 1. **Age (in years)** | | | | | |
| --- | --- | --- | --- | --- | --- |
| - 18-35 | | - 36-50 | | - Above 50 | |
| 2. **Sex** | | | | | |
| - Male | | | - Female | | |
| 3. **Education** | | | | | |
| - None | - Primary | | - Secondary | | - Higher secondary |
| - Graduate | - Postgraduate | |  | |  |
| 4. **Occupation** | | | | | |
| - Unemployed/ retired | - Student | | - Homemaker | | - Service holder |
| - Health professionals | - Daily worker | | - Business | |  |
| 5. **Residence** | | | | | |
| - Rural | | | - Urban | | |

**Section- Special**

| 1. **Have you ever heard about Mpox?** | |
| --- | --- |
| - Yes | - No |
| 2. **Where / from whom did you first hear about Mpox? (M.C.A)** | |
| - Family | - Friends or neighbors |
| - Newspaper | - Television |
| - Internet | - Social media |
| - Health workers |  |

**Section-2: Knowledge Questions**

| 1. **Mpox is prevalent in Africa.** | | | | | | | | | | | | |
| --- | --- | --- | --- | --- | --- | --- | --- | --- | --- | --- | --- | --- |
| - Yes | | | | - No | | | | | - Do not know | | | |
| 2. **There is an outbreak of human Mpox in Europe, USA, UK.** | | | | | | | | | | | | |
| - Yes | | | | - No | | | | | - Do not know | | | |
| 3. **Is there any human Mpox case found in Bangladesh?** | | | | | | | | | | | | |
| - Yes | | | | - No | | | | | - Do not know | | | |
| 4. **Mpox is a ____ disease.** | | | | | | | | | | | | |
| - Viral | | | | - Bacterial | | | | | - Do not know | | | |
| 5. **Which one/s are the carrier of Mpox virus? (M.C.A)** | | | | | | | | | | | | |
| - Squirrels | | | - Rats/mice | | | - Monkey | | | | - Do not know | | |
| 6. **What are the signs and symptoms of MPox? (M.C.A)** | | | | | | | | | | | | |
| - Fever & Headache | - Rash (Primarily) | | | - Muscle ache/myalgia | | - Swollen lymph node | | | - Skin Lesions (late-onset) | | | - Do not know |
| 7. **Mpox and smallpox have similar signs and symptoms.** | | | | | | | | | | | | |
| - Yes | | | | - No | | | | | - Do not know | | | |
| 8. **How does it transmit? (M.C.A)** | | | | | | | | | | | | |
| - Animal bite or scratch | | | | | | - Respiratory Droplets | | | | | | |
| - Direct contact with body fluids or lesion material of infected animal or human | | | | | | - Preparing/processing meat of wild animal | | | | | | |
| - Indirect contact with lesion material such as through contaminated bedding | | | | | | - Do not know | | | | | | |
| 9. **International travelers can be a source of imported cases of Mpox.** | | | | | | | | | | | | |
| - Yes | | | | - No | | | | | - Do not know | | | |
| 10. **How can you prevent MPox Virus? (M.C.A)** | | | | | | | | | | | | |
| - Avoiding contact with sick animals that can harbor the virus | | | | | | - Isolation of infected persons from others | | | | | | |
| - Avoid direct contact with materials of the infected | | | | | | - Practice good hand hygiene after contact with infected animals or humans | | | | | | |
| - Do not know | | | | | | |  | | | | | |
| 11. **There is a vaccine against the Mpox virus.** | | | | | | | | | | | | |
| - Yes | | | | - No | | | | | - Do not know | | | |
| 12. **What is/are the treatment/s for Mpox virus infection? (M.C.A)** | | | | | | | | | | | | |
| - Paracetamol | | - Antibiotic | | | - Antiviral | | | - Antihistamine | | | - Do not know | |
